# Anticoagulation trends in adults aged 65 and over with atrial fibrillation; a cohort study

**DOI:** 10.1101/2021.03.09.21253132

**Authors:** J Lund, CL Saunders, D Edwards, J Mant

## Abstract

**Objective:** To describe patterns of anticogulation prescribing and persistence for those aged ≥65 years with atrial fibrillation (AF).

**Methods:** Descriptive cohort study using electronic general practice records of patients in England who attended a flu vaccination aged ≥65, and were diagnosed with AF between 2008-2018. Patients were stratified by 10 year age group and year of diagnosis. Proportion anticoagulated, type of anticoagulation (direct oral anticoagulant (DOAC) or Warfarin) initiated at diagnosis, and persistence with anticoagulation over time are reported.

**Results:** 42,290 patients (49% female), aged 65-74 (n=11,722), 75-84 (n=19,055) and 85+ (n=11,513) at AF diagnosis are included. Prescription of anticoagulation at diagnosis increased over the time period from 55% to 86% in people aged 65-74, from 54% to 86% in people aged 75-84 and from 27% to 75% in people aged 85 and over. No patients were prescribed DOACs as a first anticoagulation agent in 2008, by 201892% of new AF patients were started on DOACs. Survivor function for 5 year persistence for patients taking only a single type of anticoagulant was 0.80 (0.77:0.82) for DOACs and0.71(0.70:0.72) for warfarin, Survivor function for any anticoagulation at 5 years was0.79(0.78:0.81), 0.73(0.72:0.75), 0.58(0.59:0.64) for people aged 65-74, 75-84 and 85+ respectively.

**Conclusions:** Rates of anticoagulation for new AF in those aged ≥65 have increased from 2008 to 2018, over which time there has been a shift from initiating anticoaguation with warfarin to DOACs. Persistence with anticoagulation is higher in people on DOACs than on warfarin, and in people under the aged of 85.

**Key Messages:** *What is already known?:* Anticoagulation is a highly effective way of reducing the risk of stroke associated with AF, but is underused, particularly in older people. The introduction of DOACs has been associated with increasing use of anticoagulation in AF.

*What does this study add?:* Our study provides up to date information on anticoagulation for AF in older people who are most at risk of AF related stroke and highlights particular increases in use of anticoagulation in people aged 85 and over. DOACs are now the major class of anticoagulant prescribed to patients with new AF in UK general practice. Long term persistence with anticoagulation is higher with DOACs than warfarin, but drops in all age groups over 5 years.

*How might this impact on clinical practice?:* Improved uptake of anticoagulation at all ages removes one of the potential barriers to screening for atrial fibrillation, but new strategies may be needed to enhance longer term persistence with treatment.

## Introduction

Atrial Fibrillation (AF) is an independent risk factor for stroke. Incidence of AF increases from 6 per 1000 in those aged 60-69 to 39 per 1000 in those aged 80-89. 31% of strokes in those aged 80-89 may be attributed to AF, compared with 7.3% in those aged 60-69.1

Oral anticoagulation reduces the risk of stroke by 65%. Though use is increasing, it is still under used in the UK, with 22% of eligible patients not receiving treatment in 2018. ^2 3^

Warfarin, a vitamin K antagonist, was the first oral anticoagulant available and was licenced in the USA in 1954. Dabigatran was the first of the Direct Oral Anticoagulation (DOAC) medications licensed for use in the UK in 2008. This has been followed by others in this group, including rivaroxaban, apixaban and edoxaban. Their arrival has been associated with an increase in prescribing of anticoagulation in non-valvular AF for all age groups.^4^

Since risk of stroke in AF rises with age, the potential benefit from anticoagulation also rises with age, though so do risk of bleeding complications.^5^ NICE guidance recommends the use of anticoagulation in all those with a CHA2DS2VASC2 score of 2 or more and the consideration of anticoagulation in those with a score of 1 where this is not due to gender, which means all patients aged ≥65 years with AF are potential candidates for anticoagulation, and all patients aged ≥75 years should be offered such treatment. Historically, older people have been less likely to receive anticoagulation despite evidence that the potential benefit is greater than the risk.^6^

AF has been considered as a candidate for a national screening programme in the UK. One of the criteria for adopting screening is that the clinical management of the condition should be optimised prior to implementation of screening. ^7^ The UK NSC (national screening committee) reviewed patterns of anticoagulation prescribing for AF in 2019. It concluded that there was insufficient evidence of optimised compliance, and around prescribing patterns of anticoagulation in AF, in part because of lack of evidence about anticoagulation persistence over time. ^8^

The objective of this study is therefore to provide this evidence of patterns of anticoagulation prescribing and persistence by 10 year age group over the age of 65.

We answered three research questions using an electronic primary care database study;

1. How has the proportion of people with AF aged over 65 who are anticoagulated changed between 2008/2018?
2. How has the type of anticoagulant prescribed changed for people with incident AF?
3. How is anticoagulation persistence over the age of 65 affected by age and type of anticoagulation?

## Method

### Data and Population

Data were extracted from the Clinical Practice Research Datalink (CPRD), a primary care research database of anonymised, electronic coded records from the Vision computer system. ^9^ All clinical activity (including diagnoses and prescribing) in primary care are recorded using clinical codes. The data-set upon which these analyses were performed was originally extracted to explore the impact of case finding for atrial fibrillation at the time of influenza vaccination. Influenza vaccination is offered to everyone over the age of 65 in the UK, and uptake in this age group over the time period of the study was 71-75%^10^.

Patients included in this study were registered with a GP in England, had records which fulfilled CPRD data quality standards^9^, had a first diagnosis of AF between 01/09/2008 and 31/08/2018, and attended at least one influenza vaccine after the age of 65 and during the study period, with at least one year’s registration before their first eligible flu vaccination. For each patient follow up started at the date of first recorded AF diagnosis and ended when the patient left the GP practice or died, the practice stopped contributing data to CPRD, or 31/08/2018, whichever was earliest. Further limited exclusions for data quality were made; patients with an AF diagnosis recorded more than 30 days after the end of follow up were excluded. Details of patient selection, inclusion and exclusion criteria are given in figure 1.

**Figure 1:**
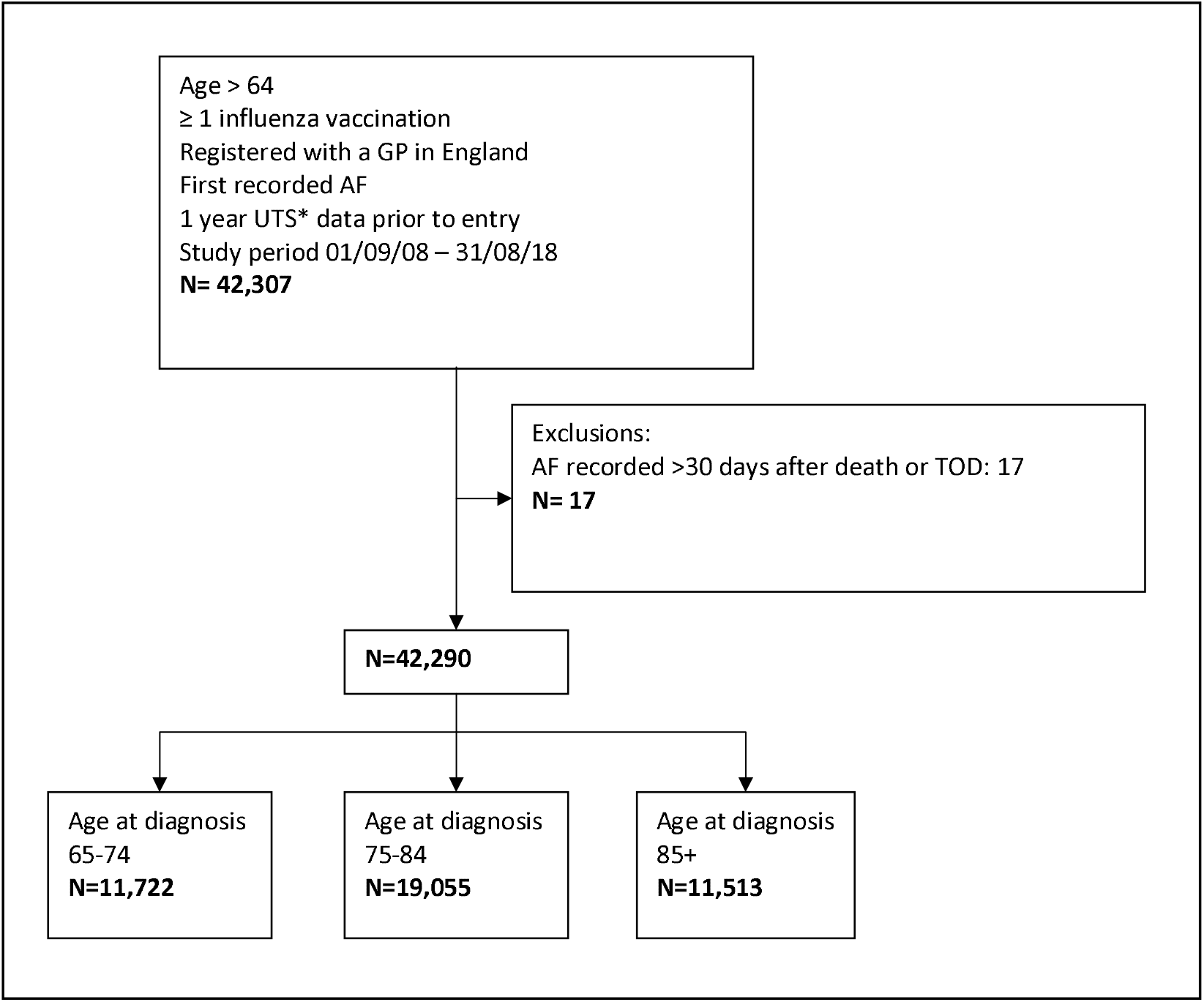
Patient flow diagram. *UTS: Up to standard. CPRD defined criteria for data quality. ^9^ TOD: Transfer out Date. Date at which patients transfers to another practice.

### Definitions of AF and anticoagulation

We analysed cases of AF diagnosed each year from 2008/09 to 2017/18 (1st September – 31 August).

The date and type of first anticoagulation are defined at the first record of an oral anticoagulant prescription. For all analyses we defined an anticoagulation prescription as a medication from the prespecified list (appendix 1) with a tablet quantity greater than zero.

Lists of codes which record AF diagnoses and anticoagulation prescribing available in supplementary data, appendix 1.

### Demographic and clinical characteristics

We defined three age groups, 65-74, 75-84 and 85+. People aged 85 and over were grouped because of small numbers and the risk of statistical disclosure. We described the clinical characteristics of the study population on the day before AF diagnosis as the count of long term conditions (out of the 20 included in the Cambridge multi morbidity score). ^11^ Deprivation was defined as the index of multiple deprivation (IMD) score of the GP practice attended by each cohort member, linked and provided by CPRD.

### Analyses

In our first analysis we explored changes in the proportion of people with AF prescribed anticoagulation by age and year. Each cohort was defined by age group at diagnosis and 2 year diagnosis interval. Patients were included in the denominator of each 2 year time point analysis if they had contributed data at any point during the 2 year time interval and were defined as anticoagulated if they had received any prescription of anticoagulation in this time.

For our second analysis we included incident AF cases who received anticoagulation within 365 days of diagnosis and calculated the proportion, with 95% confidence intervals, of these, stratified by age group, who received a DOAC as the first prescription after AF diagnosis. For this analysis patients contributed to both the numerator and denominator of any year if they had a minimum of 365 days of follow up after AF diagnosis.

In our final analyses we described the time to end of anticoagulation use, stratified by age group at diagnosis and by age and type of anticoagulation. Analysis entry point was defined as the later of AF diagnosis date or first anticoagulation prescription date. The date of end of anticoagulation is defined as the date of the last anticoagulation prescription if this is greater than 90 days before leaving the sample. If the patient left the sample for any reason within 90 days of anticoagulation prescription they were censored at the leaving date. For this analysis type of anticoagulation was defined as one of four categories; Warfarin only, DOAC only, Warfarin changed to DOAC during follow up and DOAC changed to Warfarin. Patients with a prescription for a vitamin K antagonist other than Warfarin were excluded due to small numbers. The survivor function was calculated, with 95% confidence intervals, at the 5 year time point for all age and medication cohorts.

All data analysis was completed using STATA software v.16. ^12^ There was no patient or public involvement in this project.

## Results

There were 42,290 patients with newly recorded AF during the study. Respectively 28% (11,722)), 45% (19055) and 27% (11,513) of patients were aged 65-74, 75-84 and 85+ at diagnosis. 49% (20,850) of patients in the total sample were female. The proportion of new cases diagnosed in female patients rose from 39% (4583) in the 65-74 age group to 61% (6994) in the over 85s. 35% (14861) of cases were diagnosed in those with 4 or more pre-existing comorbidities, falling to 6% (2492) in those with no other comorbidities with no systematic variation by deprivation. Across all age groups there were falling numbers of cases in the later years of the study, this represents the falling number of practices contributing to CPRD using the Vision computer software over this time period. (Table 1)

**Table 1:** Patient Characteristics.

|  |  | 08/09 | 09/10 | 10/11 | 11/12 | 12/13 | 13/14 | 14/15 | 15/16 | 16/17 | 17/18 | All years |
| --- | --- | --- | --- | --- | --- | --- | --- | --- | --- | --- | --- | --- |
| Incident AF cases |  | 4376 | 5164 | 5255 | 5576 | 5528 | 4907 | 4391 | 3172 | 2170 | 1751 | 42290 |
| By age(%) | 65 | 1204 (27.5) | 1469 (28.5) | 1410 (26.8) | 1589 (28.5) | 1548 (28.0) | 1321 (26.9) | 1192 (27.2) | 899 (28.3) | 596 (27.5) | 494 (28.2) | 11722 (27.7) |
|  | 75 | 2001 (45.7) | 2338 (45.3) | 2430 (46.2) | 2510 (45.0) | 2485 (45.0) | 2251 (45.9) | 1997 (45.5) | 1371 (43.2) | 920 (42.4) | 752 (43.0) | 19055 (45.1) |
|  | 85 | 1171 (26.8) | 1357 (26.3) | 1415 (26.9) | 1477 (26.5) | 1495 (27.0) | 1335 (27.2) | 1202 (27.4) | 902 (28.4) | 654 (30.1) | 505 (28.8) | 11513 (27.2) |
| Female(%) | 65 | 477 (39.6) | 564 (38.4) | 578 (41.0) | 627 (39.5) | 624 (40.3) | 490 (37.1) | 455 (38.2) | 355 (39.5) | 236 (39.6) | 177 (35.8) | 4583 (39.1) |
|  | 75 | 977 (48.8) | 1174 (50.2) | 1166 (48.0) | 1245 (49.6) | 1180 (47.5) | 1084 (48.2) | 1005 (50.3) | 677 (49.4) | 425 (46.2) | 340 (45.2) | 9273 (48.7) |
|  | 85 | 737 (62.9) | 827 (60.9) | 856 (60.5) | 916 (62.0) | 895 (59.9) | 831 (62.2) | 686 (57.1) | 557 (61.8) | 405 (61.9) | 284 (56.2) | 6994 (60.7) |
| IMD quintile(%)* | 1 | 864 (19.7) | 1000 (19.4) | 1023 (19.5) | 1115 (20.0) | 1019 (18.4) | 848 (17.3) | 779 (17.7) | 532 (16.8) | 410 (18.9) | 338 (19.3) | 7928 (18.8) |
|  | 2 | 773 (17.7) | 895 (17.3) | 1030 (19.6) | 1065 (19.1) | 1037 (18.8) | 944 (19.2) | 840 (19.1) | 543 (17.1) | 365 (16.8) | 278 (15.9) | 7770 (18.4) |
|  | 3 | 920 (21.0) | 1073 (20.8) | 1143 (21.8) | 1193 (21.4) | 1211 (21.9) | 1157 (23.6) | 919 (20.9) | 603 (19.0) | 415 (19.1) | 319 (18.2) | 8953 (21.2) |
|  | 4 | 873 (20.0) | 1012 (19.6) | 966 (18.4) | 949 (17.0) | 985 (17.8) | 885 (18.0) | 795 (18.1) | 579 (18.3) | 362 (16.7) | 239 (13.7) | 7645 (18.1) |
|  | 5 | 884 (20.2) | 1095 (21.2) | 989 (18.8) | 1160 (20.8) | 1165 (21.1) | 935 (19.1) | 933 (21.3) | 784 (24.7) | 495 (22.8) | 413 (23.6) | 8853 (20.9) |
| Morbidity count(%) | 0 | 283 (6.5) | 336 (6.5) | 307 (5.8) | 360 (6.5) | 318 (5.8) | 250 (5.1) | 246 (5.6) | 169 (5.3) | 113 (5.2) | 110 (6.3) | 2492 (5.9) |
|  | 1 | 767 (17.5) | 826 (16.0) | 848 (16.1) | 903 (16.2) | 875 (15.8) | 757 (15.4) | 665 (15.1) | 470 (14.8) | 300 (13.8) | 270 (15.4) | 6681 (15.8) |
|  | 2 | 992 (22.7) | 1172 (22.7) | 1200 (22.8) | 1294 (23.2) | 1185 (21.4) | 1052 (21.4) | 861 (19.6) | 627 (19.8) | 483 (22.3) | 367 (21.0) | 9233 (21.8) |
|  | 3 | 935 (21.4) | 1129 (21.9) | 1146 (21.8) | 1200 (21.5) | 1164 (21.1) | 1030 (21.0) | 920 (21.0) | 693 (21.9) | 445 (20.5) | 361 (20.6) | 9023 (21.3) |
|  | 4+ | 1399 (32.0) | 1701 (32.9) | 1754 (33.4) | 1819 (32.6) | 1986 (35.9) | 1818 (37.1) | 1699 (38.7) | 1213 (38.2) | 829 (38.2) | 643 (36.7) | 14861 (35.1) |
| Ever prescribed anticoagulation(%) | 65 | 979 (81.3) | 1243 (84.6) | 1213 (86.0) | 1369 (86.2) | 1393 (90.0) | 1280 (96.9) | 1153 (96.7) | 866 (96.3) | 573 (96.1) | 475 (96.2) | 10609 (90.5) |
|  | 75 | 1483 (74.1) | 1723 (73.7) | 1914 (78.8) | 2102 (83.7) | 2173 (87.4) | 2101 (93.3) | 1927 (96.5) | 1319 (96.2) | 900 (97.8) | 708 (94.1) | 16425 (86.2) |
|  | 85 | 413 (35.3) | 529 (39.0) | 617 (43.6) | 777 (52.6) | 879 (58.8) | 921 (69.0) | 947 (78.8) | 735 (81.5) | 553 (84.6) | 432 (85.5) | 6888 (59.8) |
| Ever prescribed DOAC(%) | 65 | 125 (10.4) | 217 (14.8) | 196 (13.9) | 255 (16.0) | 343 (22.2) | 443 (33.5) | 546 (45.8) | 588 (65.4) | 468 (78.5) | 414 (83.8) | 3660 (31.2) |
|  | 75 | 153 (7.6) | 224 (9.6) | 327 (13.5) | 404 (16.1) | 519 (20.9) | 687 (30.5) | 965 (48.3) | 929 (67.8) | 722 (78.5) | 628 (83.5) | 5633 (29.6) |
|  | 85 | 40 (3.4) | 54 (4.0) | 92 (6.5) | 132 (8.9) | 240 (16.1) | 359 (26.9) | 523 (43.5) | 538 (59.6) | 440 (67.3) | 381 (75.4) | 2884 (25.0) |
| Anticoagulation within 365 days (%) | 65 | 658 (54.7) | 773 (52.6) | 805 (57.1) | 931 (58.6) | 954 (61.6) | 951 (72.0) | 856 (71.8) | 685 (76.2) | 456 (76.5) | 382 (77.3) | 7516 (64.1) |
|  | 75 | 1075 (53.7) | 1208 (51.7) | 1301 (53.5) | 1470 (58.6) | 1576 (63.4) | 1531 (68.0) | 1432 (71.7) | 1031 (75.2) | 710 (77.2) | 579 (77.0) | 11988 (62.9) |
|  | 85 | 287 (24.5) | 359 (26.5) | 419 (29.6) | 550 (37.2) | 608 (40.7) | 682 (51.1) | 716 (59.6) | 571 (63.3) | 422 (64.5) | 358 (70.9) | 5057 (43.9) |
| DOAC within 365 days(%) | 65 | 0 (0.0) | <10 (0.0) | <10 (0.0) | 21 (1.3) | 104 (6.7) | 227 (17.2) | 378 (31.7) | 477 (53.1) | 396 (66.4) | 348 (70.4) | 2020 (17.2) |
|  | 75 | 0 (0.0) | 0 (0.0) | 0 (0.0) | 29 (1.2) | 165 (6.6) | 344 (15.3) | 683 (34.2) | 752 (54.9) | 609 (66.2) | 539 (71.7) | 3196 (16.8) |
|  | 85 | 0 (0.0) | 0 (0.0) | <10 (0.0) | 17 (1.2) | 80 (5.4) | 218 (16.3) | 394 (32.8) | 440 (48.8) | 363 (55.5) | 335 (66.3) | 1933 (16.8) |

\*Practice level IMD data is missing for 1141 patients

Both cohort and period effects are seen in the prescription of anticoagulation over the 10 years of the study. For all age groups the proportion of patients anticoagulated at the time of diagnosis increased between 2008 and 2018 (Figure 2, Table 2). This change is most marked in those aged 85+, where there was a rise from 27% (679) in 2008-2010 to 74% (865) in 2016-2018. During the same time period anticoagulation for the 65-74 age group rose from the higher baseline of 55% (1482) to 86% (937), and for the 75-84 age group from 54% (2339) to 86% (1430). The difference in the proportion of those anticoagulated between the oldest and youngest age groups narrows considerably over the 10 year period, from 28% to 11%. Within each age and year of diagnosis cohort there is also a consistent upward trend in the proportion who are anticoagulated over time. For example in those aged 85+ during 2008-2010 the proportion anticoagulated at diagnosis is 27% (679). By 2016-2018, of those who remain in this cohort, now aged 95+, 55% (56) are receiving treatment.

**Table 2:**
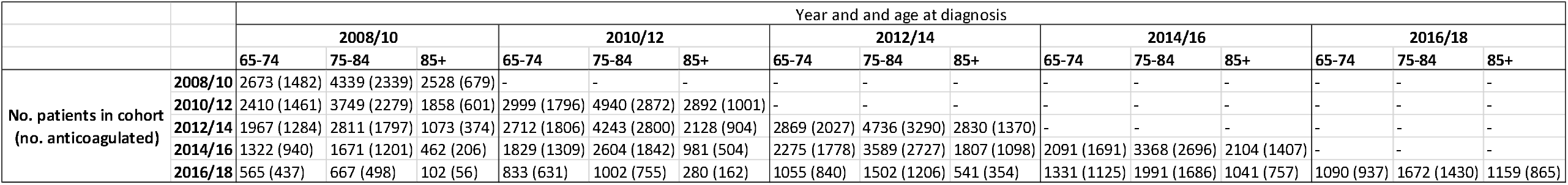
Incident cases and anticoagulation by age and year of diagnosis.

**Figure 2:**
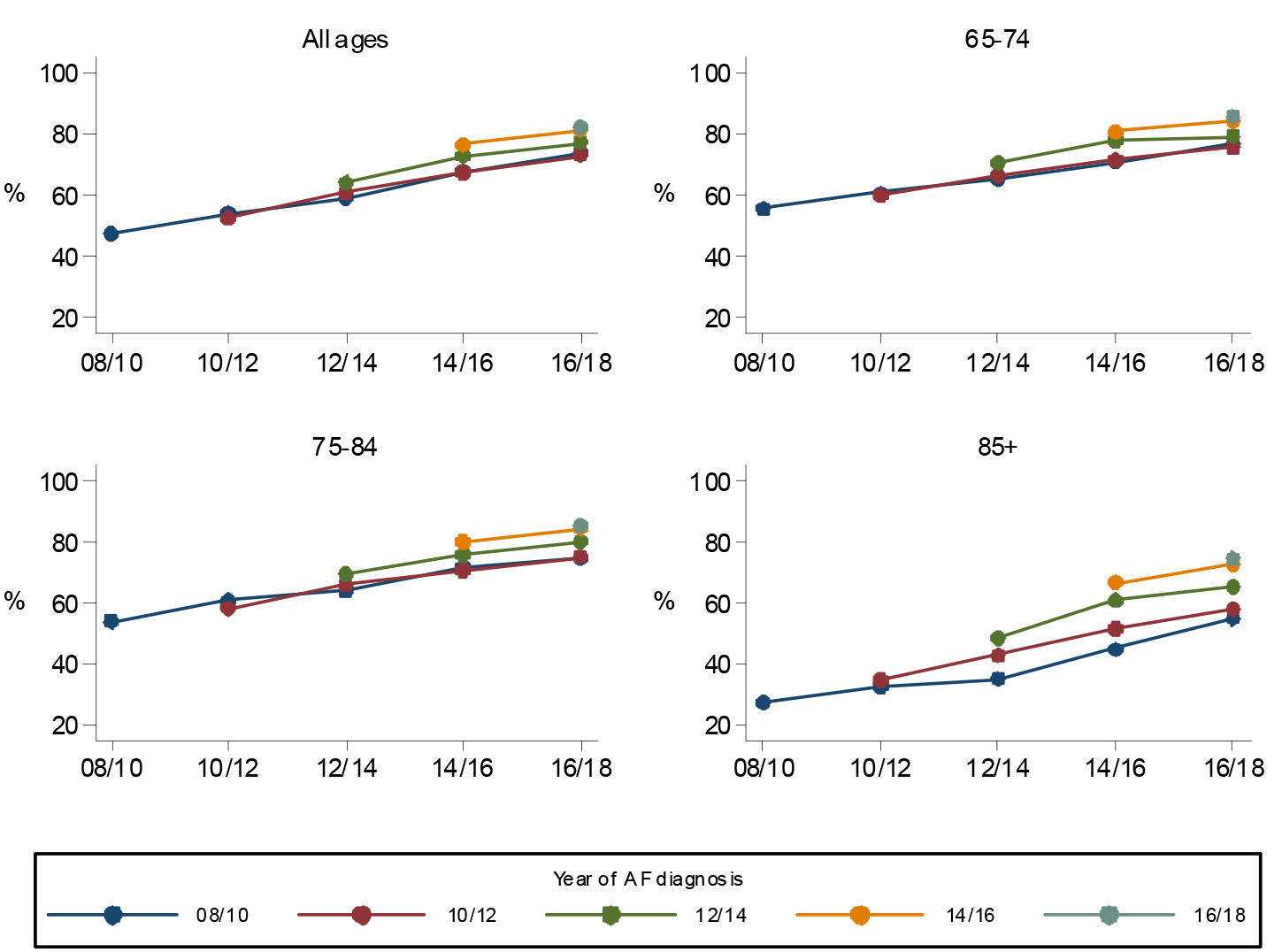
Anticoagulation for AF by cohort; age and year of diagnosis.

No patients diagnosed with AF in 2008/2009 were prescribed a DOAC, but by 2017/2018, 70% (1222) received a DOAC within 1 year of diagnosis (figure 3). DOACs represented 92% (1222/1319) of first anticoagulant prescriptions for AF in 2017/18. The use of DOACs increased rapidly in all age groups, with the largest increases seen between 2012 and 2016. The rate of increase was initially highest in the 85+ age group. There was no significant difference between age groups in the proportion given a DOAC from 2016 onwards. (Figure 3).

**Figure 3:**
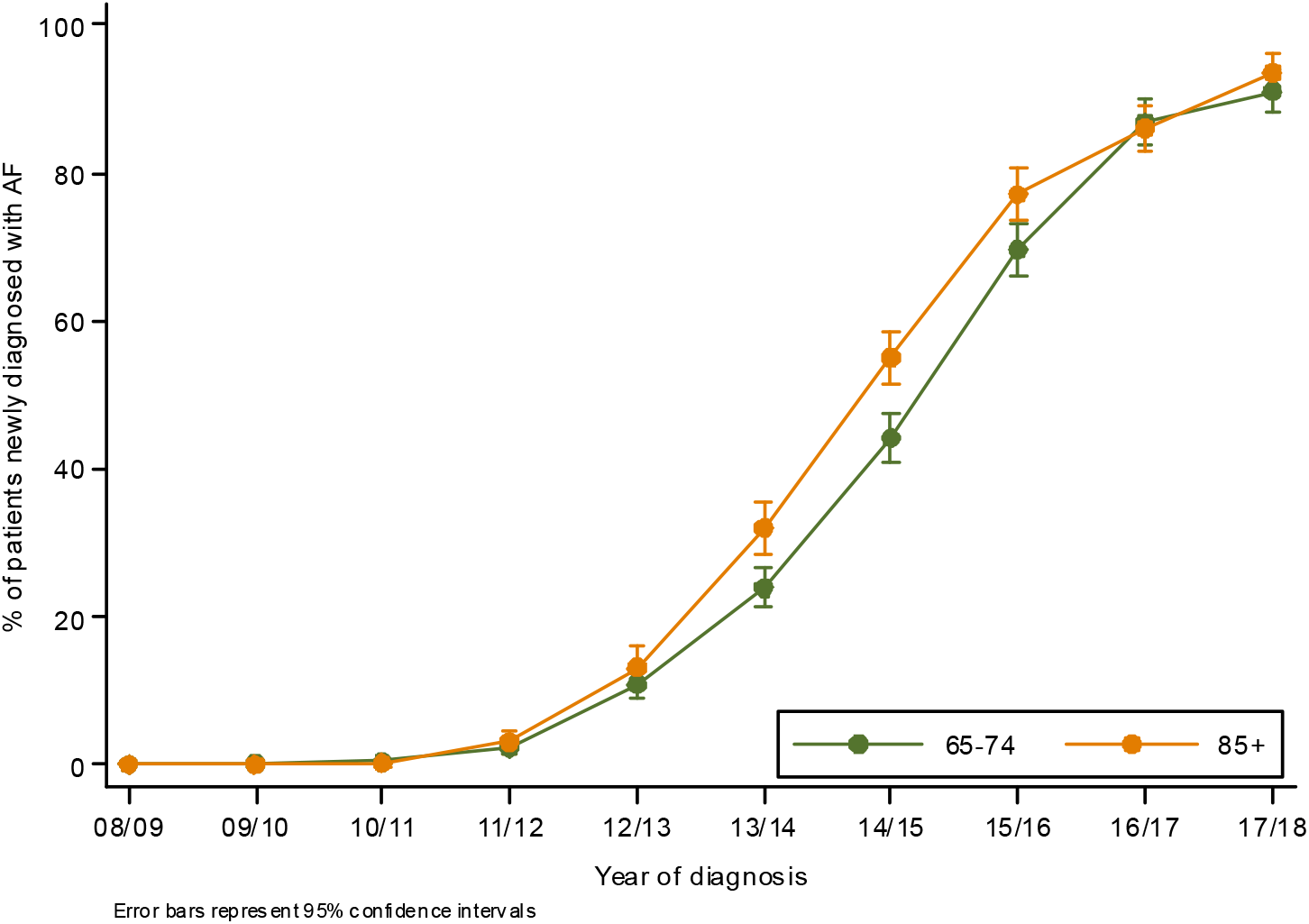
The proportion of first anticoagulation prescriptions for newly diagnosed AF that are for a DOAC.

29,644 patients received a prescription for oral anticoagulation at any timepoint following AF diagnosis. Of these 60% (17,753) received only warfarin, 28% (8351) only DOAC, 11% (3298) switched from warfarin to DOAC during the study period and 1% (246) changed from DOAC to warfarin. Due to the small number of patients who changed from DOAC to Warfarin these patients were excluded from further analysis (Table 3).

**Table 3:** Type of anticoagulation by age.

| Age at diagnosis | 65-74 | 75-84 | 85+ | All ages |
| --- | --- | --- | --- | --- |
| DOAC only | 2,379 | 3,761 | 2,211 | 8,351 |
| Warfarin only | 5,653 | 8,831 | 3,269 | 17,753 |
| Warfarin changed to DOAC | 1,125 | 1,646 | 527 | 3,298 |
| DOAC changed to Warfarin | 78 | 125 | 43 | 246 |

Persistence with anticoagulation was high across all age groups, with the greatest rate of decrease in the first year. (Figure 4) Anticoagulation persistence was higher in the younger age groups at all time points. At 5 years the survivor function for anticoagulation was 0.79 (0.78:0.81) in those aged 65-74, 0.73(0.72:0.75) in those aged 75-84 and 0.58 (0.54:0.61) in those aged 85+.

**Figure 4:**
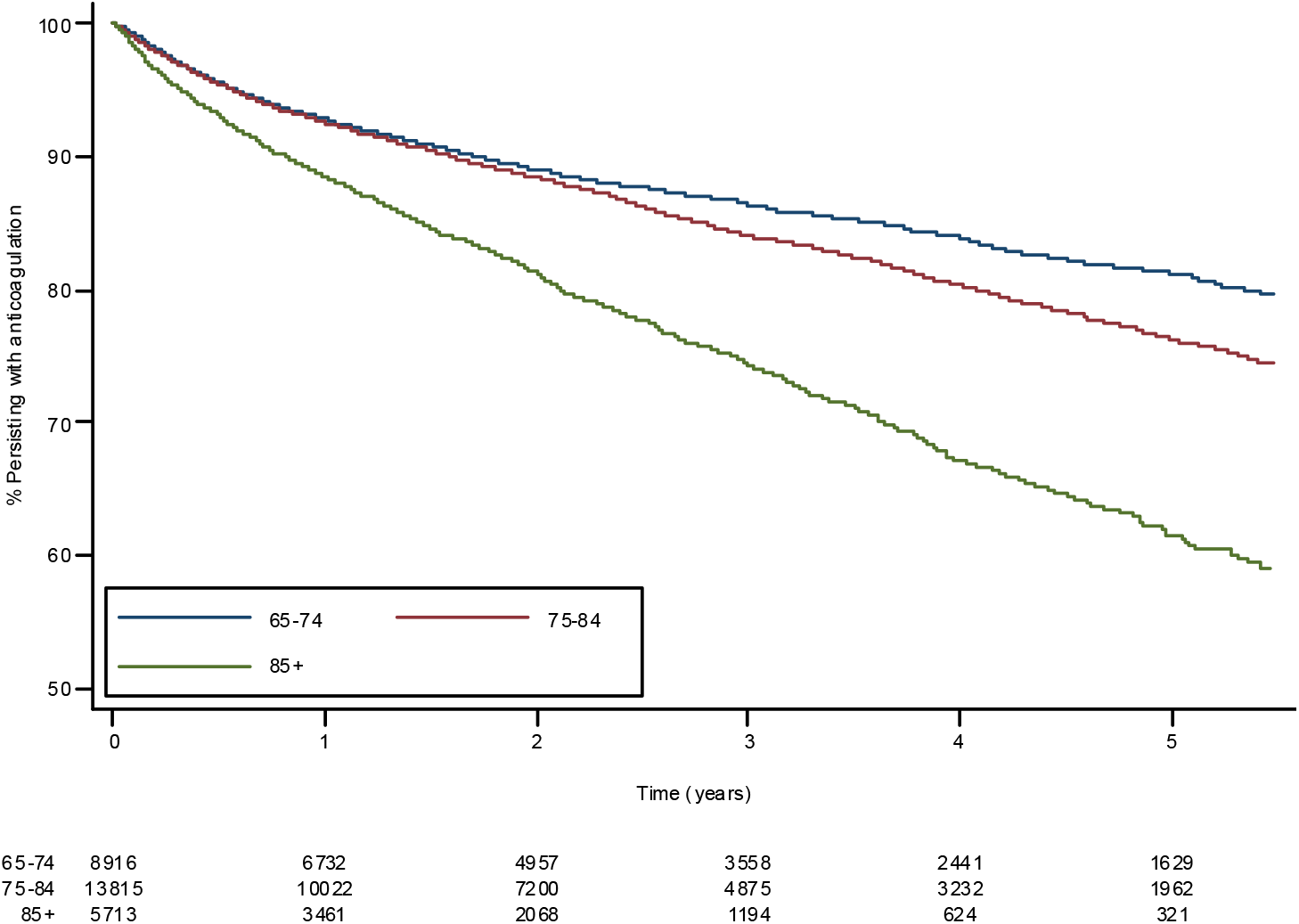
Anticoagulation persistence by age.

Of those who received a single type of anticoagulation, persistence was highest in those prescribed a DOAC (Figure 5). At 5 years the probability of continuing anticoagulation for those only prescribed a DOAC was 0.790.77:0.82) compared to 0.71(0.71:0.76) in those only prescribed Warfarin. If all patients initially prescribed warfarin are considered, including those who subsequently switched to a DOAC, then five year probability of continuing anticoagulation was 0.75 (0.74:0.76).

**Figure 5:**
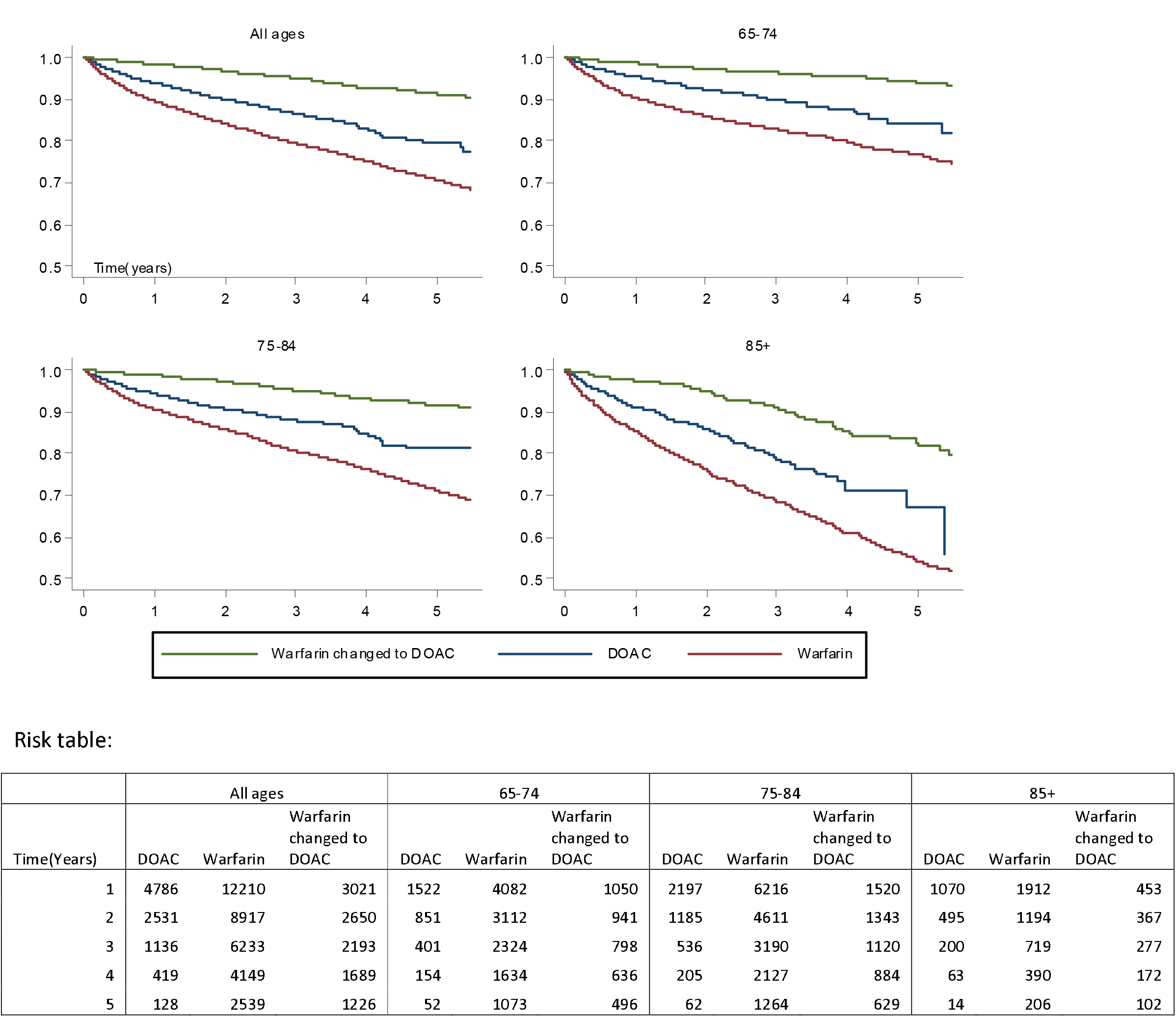
Anticoagulation persistence by age and type. Due to small patient numbers those who have transferred from DOAC to Warfarin are not presented.

## Discussion

### Summary

Anticoagulation has increased across all age groups from 2008-2018 with the greatest rise in those aged over 85. There have been both cohort and period effects with regard to prescription of anticoagulation. Increases are greatest in those with newly diagnosed AF. DOACs now represent the vast majority of anticoagulant prescriptions offered to all patients aged over 65 with newly diagnosed AF. Anticoagulation persistence decreases with age, being lowest in those aged over 85. Persistence is higher in those prescribed DOACs as compared to warfarin.

### Strengths and Limitations

The major strength of this study is the large sample size and generalisability to the UK population. Ours is the first study to tease out period from cohort effects in anticoagulation prescribing in this age group. As with all electronic record studies a major limitation is the reliance on recorded, coded, diagnoses. It is possible that patients receiving anticoagulation are more likely to have a coded diagnosis of atrial fibrillation and therefore the incidence of anticoagulation seen in this paper may be artificially elevated. Inclusion of only patients who have had an influenza vaccination may mean that the cohort of patients described in this study are those more likely to attend for healthcare and therefore more likely to receive anticoagulation. However the AF incidence seen is comparable to that reported using all CPRD records. ^13^ These potential biases are unlikely to have had any major effect on the observed trends, since their magnitude will not have changed substantially over the time period of the study.

A further limitation is the use of anticoagulant prescription issues as a marker for persistence with medication. It is possible that prescriptions are issued without medication being taken, particularly considering the move toward electronic repeat dispensing in the UK.

We have shown that anticoagulation goes up over time in all age groups. This is likely in part due to increase use of anticoagulation but may also represent differential survival in those who are anticoagulated.

### Comparison with existing literature

The trends in overall anticoagulation and the use of DOAC as the anticoagulant of choice we present are consistent with previous UK studies. ^4 14^ Our work brings these results up to date and presents new data with regard to age stratification for those aged over 65.

We show that people who were started on a DOAC tend to persist longer than those on warfarin, however there may be both patient selection and time effects. DOACs were initiated in later years than warfarin and there has been, over the period of the study, increased knowledge and awareness of the importance of persisting with anticoagulation. What we have observed is similar to the persistence seen in a recent meta-analysis where pooled persistence with DOACs was higher than vitamin K antagonists (OR 1.44), ^15^ although many of the studies included within this have a significantly shorter period of follow up. Anticoagulation persistence is known to be lower in younger age groups^16 17^, our patients were older, mean age 80 years, than in any of the studies included in the systematic review and this may partially explain the high overall persistence seen. In clinical trials where randomisation eliminates patient selection bias there was no clear pattern of superiority of DOAC over Warfarin. ^18-20^ Our study is observational and therefore we cannot be sure of the effects of patient selection bias. The choice of anticoagulant is largely an individual decision made between clinician and patient and there may be an unmeasured characteristic that affects the choice of medication and the likelihood of persistence.

### Implications for research policy and practice

Our results show there has been a major shift in the use of anticoagulation in the UK. DOACs have become the first line anticoagulant for those with AF and are being used in ever increasing numbers of patients, including large numbers aged over 85. We provide some circumstantial evidence to support the success of initiatives to increase uptake of anticoagulation. ^21^ This suggests that poor uptake of anticoagulation is no longer a barrier to screening for atrial fibrillation.

We note that the high levels of anticoagulant prescribing that we observed are against a background of multi-morbidity, which would be expected to increase the potential iatrogenic effects of anticoagulation prescribing, as multi-morbidity increases risk of haemorrhage. ^22^

Those prescribed warfarin require frequent monitoring and dose adjustment. Currently there are multiple models of care as to how this service is provided, across both primary and secondary care. As the number of patients receiving warfarin continues to decline consideration will need to be given to the best model of care for continuation of specialist warfarin services for the small number of patients who continue to use it. This will need to account for the likely older age profile of these patients.

A final consideration is that the current data are all drawn prior to the COVID-19 pandemic. As a result of the pandemic, UK guidance, endorsed by the Royal College of General Practitioners, has encouraged the accelerated switching of warfarin to DOAC for patients with AF to avoid unnecessary trips to health care settings for monitoring. ^23^ Pandemic guidance from NHS England explicitly recommends that patients newly diagnosed with AF are offered a DOAC in preference to Warfarin. Therefore the trends seen in this paper may well have accelerated in 2020.

## Conclusion

The use of anticoagulation for AF has increased over the period 2008-2018, with the greatest increases seen in those aged over 85, both in terms of initiation and persistence than previously reported. DOACs now represent the vast majority of anticoagulation prescriptions for newly diagnosed AF. This study provides some evidence that DOACs are associated with higher long term persistence in older adults than warfarin.

## Supporting information

Supplemental Tables - CPRD codelists

## Data Availability

Original data is available with license from CPRD.

https://cprd.com/home

## Acknowledgments and affiliations

Jenny Lund’s work for this study is supported by the Wellcome Trust as part of the Wellcome Trust PhD Programme for Primary Care Clinicians [grant number RHZB/195].

Catherine Saunders received salary support from the National Institute for Health Research (NIHR) School for Primary Care Research (project reference SPCR-2014-10043 Capacity Building Award 10a SPCR Postdoctoral Fellowship Award).

Jonathan Mant is an NIHR Senior Investigator.

