## Supplemental Tables - CPRD codelists for "Anticoagulation trends in adults aged 65 and over with atrial fibrillation; a cohort study"

**Appendicies**

**Appendix 1: CPRD codes defining exposure and outcomes**

**1a: Atrial Fibrillation**

| CPRD medcode | Readcode | Description |
| --- | --- | --- |
| 1268 | G573200 | Paroxysmal atrial fibrillation |
| 1664 | G573000 | Atrial fibrillation |
| 1757 | G573100 | Atrial flutter |
| 2212 | G573.00 | Atrial fibrillation and flutter |
| 23437 | G573z00 | Atrial fibrillation and flutter NOS |
| 35127 | G573300 | Non-rheumatic atrial fibrillation |
| 96076 | G573500 | Persistent atrial fibrillation |
| 96277 | G573400 | Permanent atrial fibrillation |
| 107472 | G573600 | Paroxysmal atrial flutter |

**1b: Anticoagulation**

| CPRD prodcode | Product Name | Drug substance | Dose | Anticoagulant Type |
| --- | --- | --- | --- | --- |
| 39119 | Rivaroxaban 10mg tablets | Rivaroxaban | 10mg | DOAC |
| 39444 | Dabigatran etexilate 110mg capsules | Dabigatran etexilate mesilate | 110mg | DOAC |
| 39503 | Dabigatran etexilate 75mg capsules | Dabigatran etexilate mesilate | 75mg | DOAC |
| 39639 | Xarelto 10mg tablets (Bayer Plc) | Rivaroxaban | 10mg | DOAC |
| 39755 | Pradaxa 110mg capsules (Boehringer Ingelheim Ltd) | Dabigatran etexilate mesilate | 110mg | DOAC |
| 42474 | Pradaxa 75mg capsules (Boehringer Ingelheim Ltd) | Dabigatran etexilate mesilate | 75mg | DOAC |
| 46632 | Dabigatran etexilate 150mg capsules | Dabigatran etexilate mesilate | 150mg | DOAC |
| 46678 | Pradaxa 150mg capsules (Boehringer Ingelheim Ltd) | Dabigatran etexilate mesilate | 150mg | DOAC |
| 47207 | Rivaroxaban 20mg tablets | Rivaroxaban | 20mg | DOAC |
| 47353 | Rivaroxaban 15mg tablets | Rivaroxaban | 15mg | DOAC |
| 47566 | Apixaban 2.5mg tablets | Apixaban | 2.5mg | DOAC |
| 47925 | Xarelto 20mg tablets (Bayer Plc) | Rivaroxaban | 20mg | DOAC |
| 48134 | Xarelto 15mg tablets (Bayer Plc) | Rivaroxaban | 15mg | DOAC |
| 48966 | Rivaroxaban 15mg tablets | Rivaroxaban | 15mg | DOAC |
| 53740 | Eliquis 2.5mg tablets (Bristol-Myers Squibb Pharmaceuticals Ltd) | Apixaban | 2.5mg | DOAC |
| 54066 | Apixaban 5mg tablets | Apixaban | 5mg | DOAC |
| 54451 | Rivaroxaban 20mg tablets | Rivaroxaban | 20mg | DOAC |
| 56289 | Xarelto 20mg tablets (Bayer Plc) | Rivaroxaban | 20mg | DOAC |
| 56640 | Xarelto 15mg tablets (Bayer Plc) | Rivaroxaban | 15mg | DOAC |
| 58594 | Eliquis 5mg tablets (Bristol-Myers Squibb Pharmaceuticals Ltd) | Apixaban | 5mg | DOAC |
| 62150 | Rivaroxaban 2.5mg tablets | Rivaroxaban | 2.5mg | DOAC |
| 64500 | Xarelto 2.5mg tablets (Bayer Plc) | Rivaroxaban | 2.5mg | DOAC |
| 64678 | Edoxaban 60mg tablets | Edoxaban tosilate | 60mg | DOAC |
| 65247 | Edoxaban 30mg tablets | Edoxaban tosilate | 30mg | DOAC |
| 65850 | Lixiana 60mg tablets (Daiichi Sankyo UK Ltd) | Edoxaban tosilate | 60mg | DOAC |
| 65876 | Edoxaban 15mg tablets | Edoxaban tosilate | 15mg | DOAC |
| 45 | Warfarin 1mg tablets | Warfarin sodium | 1mg | Warfarin |
| 61 | Warfarin 3mg tablets | Warfarin sodium | 3mg | Warfarin |
| 833 | Warfarin 3mg/5ml oral solution | Warfarin sodium | 600microgram/1ml | Warfarin |
| 1781 | Warfarin 5mg tablets | Warfarin sodium | 5mg | Warfarin |
| 6262 | Warfarin 500microgram tablets | Warfarin sodium | 500microgram | Warfarin |
| 8466 | Marevan 1mg tablets (AMCo) | Warfarin sodium | 1mg | Warfarin |
| 8467 | Marevan 3mg tablets (AMCo) | Warfarin sodium | 3mg | Warfarin |
| 10560 | WARFARIN 10 MG TAB |  |  | Warfarin |
| 13348 | Marevan 5mg tablets (AMCo) | Warfarin sodium | 5mg | Warfarin |
| 17965 | Marevan 500microgram tablets (AMCo) | Warfarin sodium | 500microgram | Warfarin |
| 20754 | WARFARIN |  |  | Warfarin |
| 23078 | Warfarin 1mg Tablet (WB Pharmaceuticals Ltd) | Warfarin sodium | 1mg | Warfarin |
| 30202 | Warfarin wbp 1mg Tablet (Boehringer Ingelheim Ltd) | Warfarin sodium | 1mg | Warfarin |
| 30203 | Warfarin wbp 3mg Tablet (Boehringer Ingelheim Ltd) | Warfarin sodium | 3mg | Warfarin |
| 31511 | Warfarin 3mg Tablet (WB Pharmaceuticals Ltd) | Warfarin sodium | 3mg | Warfarin |
| 31937 | Warfarin 5mg tablets (Teva UK Ltd) | Warfarin sodium | 5mg | Warfarin |
| 33711 | Warfarin 5mg Tablet (WB Pharmaceuticals Ltd) | Warfarin sodium | 5mg | Warfarin |
| 34019 | Warfarin 1mg tablets (IVAX Pharmaceuticals UK Ltd) | Warfarin sodium | 1mg | Warfarin |
| 34086 | Warfarin 3mg Tablet (Celltech Pharma Europe Ltd) | Warfarin sodium | 3mg | Warfarin |
| 34087 | Warfarin 1mg Tablet (Celltech Pharma Europe Ltd) | Warfarin sodium | 1mg | Warfarin |
| 34088 | Warfarin 5mg Tablet (Celltech Pharma Europe Ltd) | Warfarin sodium | 5mg | Warfarin |
| 34095 | Warfarin wbp 5mg Tablet (Boehringer Ingelheim Ltd) | Warfarin sodium | 5mg | Warfarin |
| 34299 | Warfarin 1mg tablets (Teva UK Ltd) | Warfarin sodium | 1mg | Warfarin |
| 34416 | Warfarin 1mg tablets (Kent Pharmaceuticals Ltd) | Warfarin sodium | 1mg | Warfarin |
| 34417 | Warfarin 3mg tablets (Teva UK Ltd) | Warfarin sodium | 3mg | Warfarin |
| 34418 | Warfarin 5mg tablets (Mylan) | Warfarin sodium | 5mg | Warfarin |
| 34517 | Warfarin 1mg tablets (Mylan) | Warfarin sodium | 1mg | Warfarin |
| 34526 | Warfarin 3mg tablets (Mylan) | Warfarin sodium | 3mg | Warfarin |
| 34576 | Warfarin 1mg Tablet (Lagap) | Warfarin sodium | 1mg | Warfarin |
| 34691 | Warfarin 5mg Tablet (Regent Laboratories Ltd) | Warfarin sodium | 5mg | Warfarin |
| 34758 | Warfarin 3mg tablets (IVAX Pharmaceuticals UK Ltd) | Warfarin sodium | 3mg | Warfarin |
| 34864 | Warfarin 5mg tablets (IVAX Pharmaceuticals UK Ltd) | Warfarin sodium | 5mg | Warfarin |
| 34918 | Warfarin 5mg tablets (Actavis UK Ltd) | Warfarin sodium | 5mg | Warfarin |
| 36099 | Warfarin 1mg/5ml oral suspension | Warfarin sodium | 200microgram/1ml | Warfarin |
| 38041 | Warfarin sodium 5mg/ml oral suspension | Warfarin Sodium | 5mg/5ml | Warfarin |
| 38044 | Warfarin 5mg/5ml oral solution | Warfarin sodium | 1mg/1ml | Warfarin |
| 39866 | Warfarin 1mg tablets (Almus Pharmaceuticals Ltd) | Warfarin sodium | 1mg | Warfarin |
| 40143 | Warfarin 500microgram tablets (A A H Pharmaceuticals Ltd) | Warfarin sodium | 500microgram | Warfarin |
| 43407 | Warfarin 3mg tablets (A A H Pharmaceuticals Ltd) | Warfarin sodium | 3mg | Warfarin |
| 43408 | Warfarin 1mg tablets (A A H Pharmaceuticals Ltd) | Warfarin sodium | 1mg | Warfarin |
| 43409 | Warfarin 5mg tablets (A A H Pharmaceuticals Ltd) | Warfarin sodium | 5mg | Warfarin |
| 43655 | Warfarin sodium oral solution | Warfarin Sodium |  | Warfarin |
| 44866 | Warfarin sodium 1mg/ml oral supension SF | Warfarin Sodium | 1mg/ml | Warfarin |
| 47944 | Warfarin 1mg tablets (Actavis UK Ltd) | Warfarin sodium | 1mg | Warfarin |
| 48070 | Warfarin sodium tablets | Warfarin Sodium |  | Warfarin |
| 48869 | Warfarin 1mg/ml oral suspension sugar free | Warfarin sodium | 1mg/1ml | Warfarin |
| 50000 | Warfarin 1mg/ml oral suspension sugar free (A A H Pharmaceuticals Ltd) | Warfarin sodium | 1mg/1ml | Warfarin |
| 51484 | Warfarin 1mg tablets (Bristol Laboratories Ltd) | Warfarin sodium | 1mg | Warfarin |
| 51496 | Warfarin 1mg tablets (Phoenix Healthcare Distribution Ltd) | Warfarin sodium | 1mg | Warfarin |
| 51509 | Warfarin 1mg tablets (APC Pharmaceuticals & Chemicals (Europe) Ltd) | Warfarin sodium | 1mg | Warfarin |
| 53745 | Warfarin 3mg tablets (Bristol Laboratories Ltd) | Warfarin sodium | 3mg | Warfarin |
| 53752 | Warfarin 1mg tablets (Alliance Healthcare (Distribution) Ltd) | Warfarin sodium | 1mg | Warfarin |
| 54892 | Warfarin 1mg/ml oral suspension sugar free (Alliance Healthcare (Distribution) Ltd) | Warfarin sodium | 1mg/1ml | Warfarin |
| 54946 | Warfarin 3mg tablets (Actavis UK Ltd) | Warfarin sodium | 3mg | Warfarin |
| 55316 | Warfarin 3mg/5ml oral suspension | Warfarin sodium | 600microgram/1ml | Warfarin |
| 56314 | Warfarin 3mg tablets (Kent Pharmaceuticals Ltd) | Warfarin sodium | 3mg | Warfarin |
| 57032 | Warfarin 1mg/ml oral suspension sugar free (Rosemont Pharmaceuticals Ltd) | Warfarin sodium | 1mg/1ml | Warfarin |
| 58519 | Warfarin 1mg tablets (DE Pharmaceuticals) | Warfarin sodium | 1mg | Warfarin |
| 58787 | Warfarin 5mg tablets (Alliance Healthcare (Distribution) Ltd) | Warfarin sodium | 5mg | Warfarin |
| 58962 | Warfarin 3mg tablets (DE Pharmaceuticals) | Warfarin sodium | 3mg | Warfarin |
| 59400 | Warfarin 500microgram tablets (Sigma Pharmaceuticals Plc) | Warfarin sodium | 500microgram | Warfarin |
| 59578 | Warfarin 3mg tablets (Phoenix Healthcare Distribution Ltd) | Warfarin sodium | 3mg | Warfarin |
| 60589 | Warfarin 500microgram tablets (Actavis UK Ltd) | Warfarin sodium | 500microgram | Warfarin |
| 60949 | Warfarin 5mg/5ml oral suspension | Warfarin sodium | 1mg/1ml | Warfarin |
| 62309 | Warfarin 500microgram tablets (Kent Pharmaceuticals Ltd) | Warfarin sodium | 500microgram | Warfarin |
| 62310 | Warfarin 500microgram tablets (AMCo) | Warfarin sodium | 500microgram | Warfarin |
| 63071 | Warfarin 4mg tablets | Warfarin sodium | 4mg | Warfarin |
| 65285 | Warfarin 1mg tablets (Crescent Pharma Ltd) | Warfarin sodium | 1mg | Warfarin |
| 65496 | Warfarin 500microgram tablets (Phoenix Healthcare Distribution Ltd) | Warfarin sodium | 500microgram | Warfarin |
| 65746 | Warfarin 500microgram tablets (DE Pharmaceuticals) | Warfarin sodium | 500microgram | Warfarin |
